# Cross-neutralization of Bundibugyo virus following Ebola virus booster vaccination

**DOI:** 10.64898/2026.11.23.26368389

**Authors:** Christian M. Kahusu, Selien Oostvogels, Johan Michiels, Emilie Dhondt, Magali Buys, Achilleas Tsoumanis, EBO-BOOST-BRIDGE consortium, Jacques Likofata, Daniel Mukadi-Bamuleka, Hugo Kavunga-Membo, Sabue Mulangu, Kevin K. Ariën, Wim Adriaensen, the EBO-BOOST-BRIDGE consortium

## Abstract

**Background:** The ongoing Bundibugyo virus disease (BVD) outbreak in the Democratic Republic of Congo (DRC) highlights the absence of a licensed Bundibugyo virus (BDBV) vaccine. Ebola virus (EBOV) vaccines can induce cross-reactive BDBV binding antibodies, but evidence of cross-neutralizing antibodies is limited.

**Methods:** In the EBO-BOOST trial (NCT06126822), previously EBOV-vaccinated adults received an ERVEBO® booster after primary vaccination with ERVEBO® (E|E, n=60) or ZABDENO®/MVABEA® (Zm|E, n=40). Neutralizing antibodies against EBOV and BDBV were measured at baseline, day 21 (D21) and month 6 (M6) using biologically contained EBOV-ΔVP30-eGFP and BDBV-ΔVP30-eGFP reporter viruses.

**Results:** BDBV seropositivity increased from 8% at baseline to 67% at D21, declining to 34% at M6. Median NT50 titers were significantly lower for BDBV than for EBOV at D21 (40.9 vs 244.1; P<0.001) and at M6 (<20 vs 66.5; P<0.001). In mixed-effects models adjusted for time since primary vaccination, BDBV responses were 65% lower than EBOV responses at D21 (GMR 0.35; 95% CI 0.26–0.48) and 50% lower at M6 (GMR 0.50; 95% CI 0.40–0.63). The Zm|E group showed a higher BDBV fold change from baseline at D21 than the E|E group (GMR 1.75; p=0.023).

**Conclusions:** An ERVEBO® booster induces BDBV cross-neutralizing antibodies in most recipients, but responses are lower than against EBOV and largely wane by 6 months. Although protective significance and correlates of protection remain to be established, these findings support evaluation of ERVEBO® in parallel with BDBV-specific vaccines.

## Letter to the editor (correspondence)

The ongoing outbreak of Bundibugyo virus disease in the Democratic Republic of Congo (DRC) highlights the lack of preventive vaccination against Bundibugyo virus (BDBV). Ebola virus (EBOV) vaccines elicit cross-reactive BDBV binding antibodies, albeit at 2.8-fold lower levels than against EBOV^1,2^. Modelling efforts suggest this to remain above putative protective thresholds, if we assume these are reached between 7 to 10 days after vaccination^3^. The evidence for BDBV cross-neutralization however is limited to a study of 10 ERVEBO® vaccinated Phase I participants using a pseudovirus assay that showed a 3.5 times less efficient neutralization^2^, and therefore remains incomplete to inform policy.

In the EBO-BOOST trial (NCT06126822), previously EBOV vaccinated adults in DRC received an ERVEBO® booster following primary vaccination with ERVEBO® or ZABDENO/MVABEA®, referred to as E|E and Zm|E respectively. Serum was collected at baseline (D0, pre-boost), day 21 (D21), and month 6 (M6) post-boost. Neutralizing antibody responses against EBOV and BDBV were assessed for 40 unvaccinated participants (unexposed group from Kinshasa), 60 participants in the E|E arm and 40 participants in the Zm|E arm. We used a biologically contained EBOV-ΔVP30-eGFP and BDBV-ΔVP30-eGFP reporter virus platform^4,5^ (**Appendix**).

For EBOV, seropositivity increased from 25% (25/100) at baseline to 90% at D21, before declining to 83% at M6. Corresponding rates for BDBV were 8%, 67% and 34%, respectively (**Appendix**). The median 50% neutralizing titre (NT_50_) value at baseline was <20 for both species, but was significantly lower for BDBV than for EBOV at D21 (40.89 vs. 244.14; P<0.001) and M6 (<20 vs. 66.46; P<0.001) **(Figure 1A)**. mIU/mL and NT90 values provided in **Appendix**. In mixed-effects regression models adjusted for time since primary vaccination, BDBV neutralization responses were 65% lower than EBOV responses at D21 (GMR, 0.35; 95% CI, 0.26-0.48) and 50% lower at M6 (GMR, 0.50; 95% CI, 0.40-0.63). For BDBV, the Zm|E group showed a higher fold change at D21 from baseline than the E|E group (1.75 GMR, p=0.023), and a borderline non-significant GMR fold-change of 1.31 at M6 (p=0.055) (**Figure 1B**), suggesting an impact of the heterologous primary vaccination regimen. Commonly, BDBV-neutralizing antibody responses following an ERVEBO® booster are present to some extend but short-lived in most individuals. Given the long interval since primary vaccination, comparable or slightly lower responses may be expected following primary vaccination alone. Although protective significance and correlates of protection remain to be established, these findings support evaluation of ERVEBO® in parallel with BDBV-specific vaccines.

**Figure 1:**
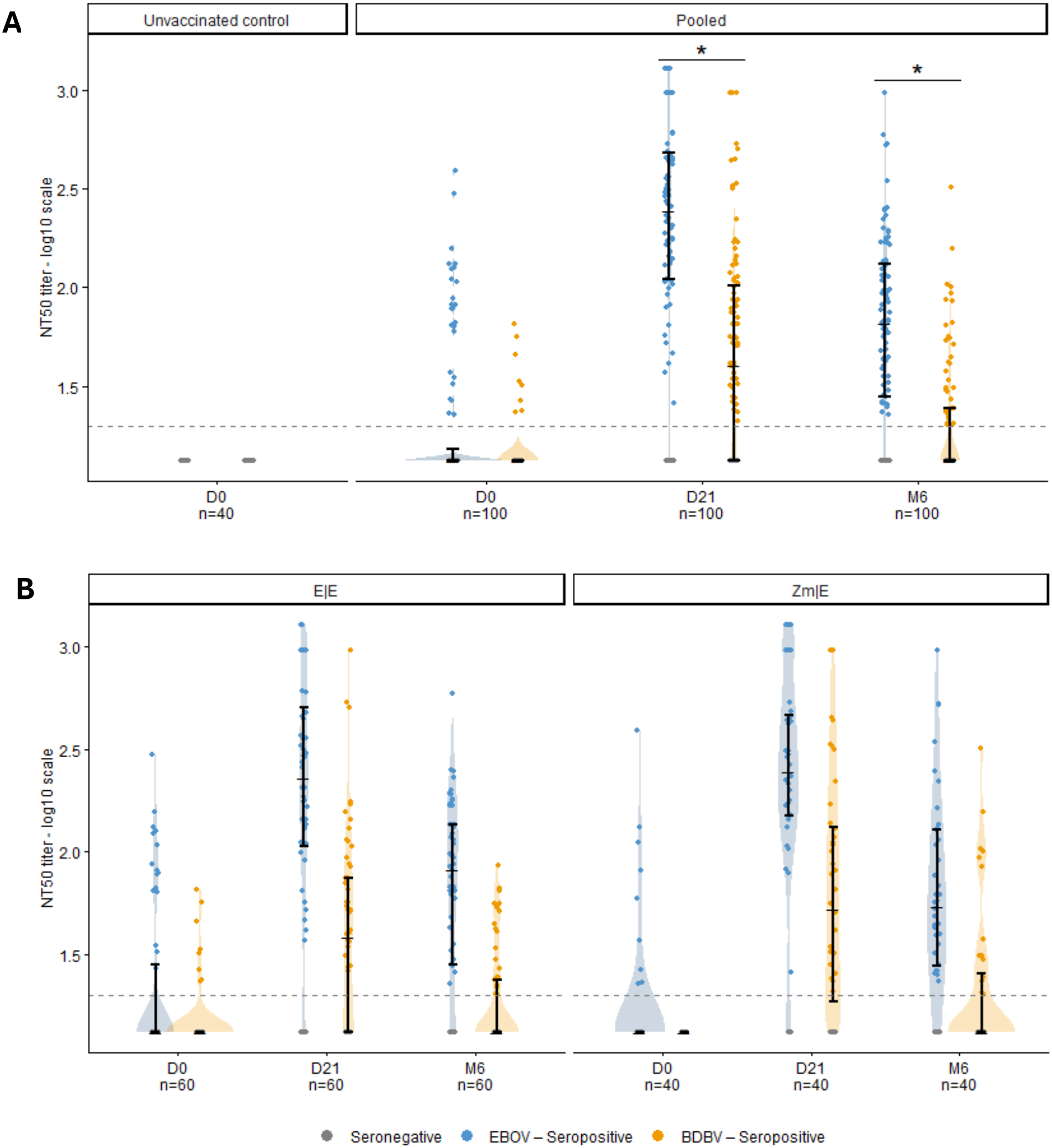
Violin plots showing the distribution of half-maximal neutralization titres (NT50) of EBOV (blue) and BDBV (orange) with medians and interquartile ranges (IQR) for (A) the unvaccinated control group and all vaccinated participants (referred to as ‘pooled’), and (B) stratified for participants having received the Ervebo|Ervebo (E/E) vaccination schedule or the Zabdeno-Mvabea/Ervebo (Zm|E) vaccination schedule. The dashed lines in grey represents the seropositivity threshold, set to the lowest limit of detection (observed in all unvaccinated controls). Pairwise comparisons of geometric mean responses between EBOV and BDBV were performed at each timepoint using paired t-tests with Benjamini-Hochberg correction for multiple comparisons. All comparisons were statistically significant (adjusted p < 0.01). * indicates a significant difference between EBOV and BDBV geometric mean ratio (GMR) in a mixed effects regression model with adjustment for time since primary vaccination (p < 0.05).

## Data Availability

De-identified participant data will be available upon reasonable request to the corresponding author following publication, subject to approval by the study Data Access Committee and applicable ethical and regulatory requirements.

## Acknowledgments

We are grateful to all members of the Laboratoire Provincial in Mbandaka, the Programme Elargi de Vaccination (PEV) in Mbandaka for the use of their facilities, and to all consented participants. In particular we like to thank Drs. Jens Verlinden, Bert Vanmechelen and Kurt Vermeire for providing the ΔVP30-eGFP reporter virus system.

## Funder statement

This study was funded by Coalition for Epidemic Preparedness Innovations (CEPI), MSD, and the Belgian Directorate-General for Development Cooperation and Humanitarian Aid (DGD).

## Notes

### Competing Interest Statement

The authors have declared no competing interest.

### Clinical Trial

NCT06126822

### Author Declarations

Ethics committee/IRB of Institute of Tropical Medicine (ITM) gave ethical approval for this work. Ethics committee/IRB of Unversitair Ziekenhuis Antwerpen (UZA) gave ethical approval for this work. Ethics committee/IRB of Comite National d'Ethique de la Sante (CNES) gave ethical approval for this work. Regulatory Authority/RA of Authorite Congolaise de Reglementation Pharmaceutique (ACOREP) gave ethical approval for this work.

