## Appendix for "Cross-neutralization of Bundibugyo virus following Ebola virus booster vaccination"

*Participant characteristics*

|  | **Unvaccinated control**  **(n = 40)** | **Total vaccinated**  **(n = 100)** | **E\|E**  **(n = 60)** | **Zm\|E**  **(n = 40)** | **Adjusted p-value for Zm\|E and E\|E comparison** |
| --- | --- | --- | --- | --- | --- |
| Basic demographics | | | | | |
| Age (years), median (IQR) | 26.0 (23.0 – 35.0) | 32.0 (24.8 – 42.0) | 31.5 (25.0 – 41.2) | 33.5 (22.0 – 42.2) | *NA* |
| Sex (female), n (%) | 19 (47.5) | 55 (55.0) | 30 (50) | 25 (62.5) | *NA* |
| Year of primary vaccination, n (%) | | | | | |
| 2018 | *NA* | 20 (20.0) | 20 (33.3) | 0 (0) | *NA* |
| 2020 | *NA* | 20 (20.0) | 20 (33.3) | 0 (0) | *NA* |
| 2022 | *NA* | 20 (20.0) | 20 (33.3) | 0 (0) | *NA* |
| 2023 | *NA* | 40 (40.0) | 0 (0) | 40 (100) | *NA* |
| NT50 EBOV, median (IQR) | | | | | |
| D0 | 13.3 (13.3–13.3) | 13.3 (13.3–15.7) | 13.3 (13.3–28.4) | 13.3 (13.3–13.3) | 0.66 |
| D21 | *NA* | 244.1 (109.9–476.4) | 231.4 (105.9–505.3) | 247.8 (149.0–460.6) | 0.66 |
| M6 | *NA* | 66.5 (28.0–132.3) | 82.9 (28.0–135.1) | 54.5 (27.9–128.5) | 0.92 |
| NT50 EBOV seropositivity, n (%) | | | | | |
| D0 | 0 | 25 (25.0) | 16 (26.7) | 9 (22.5) | 0.81 |
| D21 | *NA* | 90 (90.0) | 53 (88.3) | 37 (92.5) | 0.81 |
| M6 | *NA* | 83 (83.0) | 48 (80.0) | 35 (87.5) | 0.81 |
| NT50 BDBV, median (IQR) | | | | | |
| D0 | 13.3 (13.3–13.3) | 13.3 (13.3–13.3) | 13.3 (13.3–13.3) | 13.3 (13.3–13.3) | 0.021* |
| D21 | *NA* | 40.1 (13.3–101.6) | 38.5 (13.3–74.5) | 53.3 (19.1–132.0) | 0.13 |
| M6 | *NA* | 13.3 (13.3–24.4) | 13.3 (13.3–23.9) | 13.3 (13.3–25.8) | 0.35 |
| NT50 BDBV seropositivity, n (%) | | | | | |
| D0 | 0 | 8 (8.0) | 8 (13.3) | 0 | 0.061 |
| D21 | *NA* | 67 (67.0) | 37 (61.7) | 30 (75.0) | 0.29 |
| M6 | *NA* | 34 (34.0) | 20 (33.3) | 14 (35.0) | 1.00 |
| NT90 EBOV, median (IQR) | | | | | |
| D0 | 10.0 (10.0–10.0) | 10.0 (10.0–10.8) | 10.0 (10.0–13.3) | 10.0 (10.0–10.0) | 0.77 |
| D21 | *NA* | 100.2 (51.2–221.3) | 94.3 (45.6–215.2) | 119.4 (69.3–230.1) | 0.76 |
| M6 | *NA* | 28.6 (13.3–57.2) | 32.1 (13.3–58.5) | 23.8 (13.3–53.9) | 0.79 |
| NT90 EBOV seropositivity, n (%) | | | | | |
| D0 | 0 | 17 (17.0) | 12 (20.0) | 5 (12.5) | 0.83 |
| D21 | *NA* | 87 (87.0) | 51 (85.0) | 36 (90.0) | 0.83 |
| M6 | *NA* | 69 (69.0) | 42 (70.0) | 27 (67.5) | 0.83 |
| NT90 BDBV, median (IQR) | | | | | |
| D0 | 10.0 (10.0–10.0) | 10.0 (10.0–10.0) | 10.0 (10.0–10.0) | 10.0 (10.0–10.0) | 0.069 |
| D21 | *NA* | 20.5 (10.0–42.5) | 13.3 (10.0–26.2) | 22.7 (12.5–57.8) | 0.076 |
| M6 | *NA* | 10.0 (10.0–13.3) | 10.0 (10.0–13.3) | 10.0 (10.0–13.3) | 0.16 |
| NT90 BDBV seropositivity, n (%) | | | | | |
| D0 | 0 | 2 (2.0) | 2 (3.3) | 0 | 0.55 |
| D21 | *NA* | 51 (51.0) | 29 (48.3) | 22 (55.0) | 0.55 |
| M6 | *NA* | 12 (12.0) | 6 (10.0) | 6 (15.0) | 0.55 |
| NT50 EBOV in mIU/mL, median (IQR) | | | | | |
| D0 | 88.7 (88.7-88.7) | 88.7 (88.7-104.8) | 88.7 (88.7 – 189.0) | 88.7 (88.7-88.7) | 0.70 |
| D21 | *NA* | 1623.5 (731.3-3167.8) | 1538.5 (704.5– 3359.5) | 1647.5 (991.3–3062.8) | 0.27 |
| M6 | *NA* | 442.5 (186.8-880.0) | 551.5 (186.8–898.3) | 363.0 (185.8–855.0) | 0.27 |
| NT50 BDBV in mIU/mL, median (IQR) | | | | | |
| D0 | 88.7 (88.7–88.7) | 88.7 (88.7–88.7) | 88.7 (88.7 – 88.7) | 88.7 (88.7–88.7) | 0.021* |
| D21 | *NA* | 272.0 (88.7–674.3) | 256.0 (88.7 – 495.0) | 354.0 (127.2–876.3) | 0.61 |
| M6 | *NA* | 88.7 (88.7–162.3) | 88.7 (88.7–159.0) | 88.7 (88.7–171.8) | 0.45 |

*Supplementary table 1: Overview of basic demographic participant data including NT50, NT50 converted to mIU/mL, NT90 and seropositivity against Ebola virus (EBOV) and Bundibugyo virus (BDBV). P-values for NT50, NT90 and mIU/mL titers represent comparisons between the E|E and Zm|E groups at each species and timepoint using unpaired t-tests on long10-transformed values. P-values for seropositivity are calculated using Fisher-s exact tests. All p-values were adjusted for multiple comparisons using the Benjamini-Hochberg correction; * indicates adjusted p < 0.05. Abbreviations: BDBV, Bundibugyo virus; EBOV, Ebola virus, IQR, interquartile range; mIU/mL, mili-international units per mililiter; NA, not applicable, NT50, 50% neutralization titer; NT90, 90% neutralization titer, neutralization titer 90 (NT90), and corresponding seropositivity values against Ebola virus (EBOV) and Bundibugyo virus (BDBV).*

*EBOV-ΔVP30-eGFP and BDBV-ΔVP30-eGFP reporter virus platform details***:**

Neutralising antibody titres were assessed using vp30-deficient, biologically contained EBOV (Mayinga strain) and BDBV (Butalya 2007 strain) carrying an eGFP-encoding reporter gene and Vero E6 cell lines expressing the VP30 protein of the respective virus. Two-fold dilutions (1/20-1/640) of heat-inactivated serum samples in assay medium were preincubated in the presence of 3 TCID100 of virus per 100 µl for 1h at 37°C / 7% CO_2_. Virus, cell and sample controls in assay medium were incubated in parallel. Subsequently, 100 µl of sample-virus mixture and controls was added to 20.000 cells in 100 µl of assay medium per well in a 96-well cell culture plate and incubated at 37°C / 7% CO_2_. All sample dilutions were tested in 8 replicates. After 6 days (for EBOV) or 7 days (for BDBV) of culture, viral replication was quantified by measuring eGFP fluorescence using a Tristar LB941 luminometer (Berthold Technologies). 50% (NT50) and 90% (NT90) neutralisation titres were calculated from the relative reduction in fluorescence intensity compared with the virus control. NT50 results were additionally expressed as mIU/ml using an internal standard consisting of pooled samples from vaccinated participants in the study that was calibrated against the WHO 1^st^ International Standard for EBOV antibodies (NIBSC code 15/262; conversion rate for EBOV: 5119mIU/ml for NT50 = 770; for BDBV: 823 mIU/mL for NT50 = 124). The seropositivity threshold was defined as the median plus three times the standard deviation of the unvaccinated group, which corresponded to the lower limit of detection of the assay.

EBO-BOOST-BRIDGE consortium

Wim Adriaensen^1^ M.Sc. Ph.D., Steve Ahuka^2,3^ M.D., Ph.D.,Diana Arango^1^ M.Sc., Ahidjo Ayouba^4^ M.Sc., Ph.D., Ella Baert^1^ M.Sc., Lucien Boba^5^, Magali Buys^6^ M.Sc., Ann Ceulemans^6^ B.Sc., Emilie Dhondt^6^ B.Sc., Thomas Hendrickx^1^ M.Sc., Christian. M. Kahusu^1,5^ M.Sc., M.D., Hugo Kavunga-Membo^7,8^ M.D., Laurens Liesenborghs^1^ M.D., Ph.D., Jacques Likofata^9^ M.Sc., Joachim Mariën^6^ M.Sc., Ph.D., Placide Mbala^2,10^ M.D., Ph.D., Johan Michiels^6^ B.Sc., Daniel Mukadi-Bamuleka M.Sc.^3,7,11^, Ph.D., Saidou Milua Lusala^5^ B.Sc., Myriam Mbili Ilombe^5^ B.Sc., Kristel Mertens^1^ M. Sc., Kevin. K. Ariën^6,12^ Ph.D, Sabue Mulangu^5,12,13^ M.D., Selien Oostvogels^1^ M.Sc., Martine Peeters^4^ M.Sc., Ph.D., Lindsay Poppe^1^ M. Sc., Ph.D., Jessy Sabue^5^ M.D., Aliana Saenz de la Torre Leal^1^ B.Sc., M.Sc., Samuel Shamamba^5^ M.D., Patrick Tshita^5^ M.Sc., Achilleas Tsoumanis^1^ M.Sc., Ph.D., Johan Van Griensven^1^ M.D., Ph.D., Yven Van Herrewege^1^ M.Sc., Ph.D., Harry Van Loen^1^ M.Sc.

1 Department of Clinical Sciences, Institute of Tropical Medicine (ITM), Antwerp, Belgium

2 Department of Medical Biology, Faculty of Medicine, University of Kinshasa (UNIKIN), DRC

3 Virology Department, Institut National de Recherche Biomédicale (INRB), Kinshasa, DRC

4 Recherche Translationnelle Appliquée au VIH et aux Maladies Infectieuses, University of Montpellier, Institut de Recherche pour le Développement, Montpellier, France

5 Immunology Department, Institut National de Recherche Biomédicale (INRB), Kinshasa, DRC

6 Department of Biomedical Sciences, Institute of Tropical Medicine (ITM), Antwerp, Belgium

7 Rodolphe Mérieux Institut National de Recherche Biomédicale (INRB) Goma, Goma, North Kivu, DRC

8 Vaccinology Department, Institut National de Recherche Biomédicale (INRB), Kinshasa, DRC

9 Laboratoire Provincial, Mbandaka, Equateur, DRC

10 Epidemiology and Global Health, Institut National de Recherche Biomédicale (INRB), Kinshasa, DRC

11 Service of Microbiology, Department of Medical Biology, Faculty of Medicine, University of Kinshasa, DRC

12 Department of Biomedical Sciences, University of Antwerp, Antwerp, Belgium

13 Ridgeback Biotherapeutics, Miami, FL, USA
